# Determination of Robust Regional CT Radiomics Features for COVID-19

**DOI:** 10.1101/2020.06.24.20139410

**Authors:** Mahbubunnabi Tamal

## Abstract

**Background:** The lung CT images of COVID-19 patients can be characterized by three different regions – Ground Glass Opacity (GGO), consolidation and pleural effusion. GCOs have been shown to precede consolidations. Quantitative characterization of these regions using radiomics can facilitate accurate diagnosis, disease progression and response to treatment. However, according to the knowledge of the author, regional CT radiomics analysis of COVID-19 patients has not been carried out. This study aims to address these by determining the radiomics features that can characterize each of the regions separately and can distinguish the regions from each other.

**Methods:** 44 radiomics features were generated with four quantization levels for 23 CT slice of 17 patients. Two approaches were the implemented to determine the features that can differentiate between lung regions – 1) Z-score and correlation heatmaps and 2) one way ANOVA for finding statistically significantly difference (p<0.05) between the regions. Radiomics features that show agreement for all cases (Z-score, correlation and statistical significant test) were selected as suitable features. The features were then tested on 52 CT images.

**Results:** 10 radiomics features were found to be the most suitable among 44 features. When applied on the test images, they can differentiate between GCO, consolidation and pleural effusion successfully and the difference provided by these 10 features between three lung regions are statistically significant.

**Conclusion:** The ten robust radiomics features can be useful in extracting quantitative data from CT lung images to characterize the disease in the patient, which in turn can help in more accurate diagnosis, staging the severity of the disease and allow the clinician to plan for more successful personalized treatment for COVID-19 patients. They can also be used for monitoring the progression of COVID-19 and response to therapy for clinical trials.

## INTRODUCTION

COVID-19 caused by severe acute respiratory syndrome coronavirus 2 (SARS-COV-2) is the worldwide leading cause of death at the moment and is a public health emergency concern across the globe [1]. A total of 437,532 deaths with a confirmed infection 8,063,488 has been reported [2].

To control and minimize the spread of COVID-19, it is important to diagnose it as quickly and accurately as possible. Though reverse transcription polymerase chain reaction (RT-PCR) is considered as a gold standard point-of-care diagnostic tool with the highest specificity [3], it requires several hours to get the results, the sensitivity is very low with a very high rate of false negatives and cannot provide the detail condition of the patient [4].

On the other hand, it has been reported that 20% people who are suffering from COVID-19 needs to visit hospital because of respiratory distress and 30% of them require intensive care [5]. In the absence of appropriate drug, two third of the COVID-19 patients who goes to critical care requires prolong period of mechanical ventilation within 24 hours of admission [6]. This group of patients also requires continuous monitoring of their lung health to assess their overall conditions.

Because of its high sensitivity specially within the first two days of infection and capability to provide detail condition of the patient, computed tomography (CT) is routinely being used as a diagnostic and screening tool in several countries including China [7-11]. Quantitative imaging information such as the lesion percentage and the CT mean density values could help the physician to monitor the progression of the disease and the response to therapy [12]. More in depth analysis of different first, second and third order statistical CT features known as radiomics have been successfully utilized for decoding the radiographic phenotype in cancer [13, 14]. Accurate quantification of tumour radiomic features was made possible by utilizing AI [15]. In case of COVID-19, only a very few studies have been carried out for radiomics analysis. One of the studies has shown that CT radiomics not only can help to screen patients [16] but also can be used as a discriminating tool for diagnosis and monitoring different stages of respiratory abnormalities [17]. Utilizing AI with radiomics can make the process faster and more accurate [18].

All the studies so far extracted the radiomics features from the whole CT lung region [16, 18, 19]. The lung on a CT of a COVID-19 patient can contain either one or any combinations of three different regions - ground-glass opacities (GCO), consolidation and pleural effusion as shown in Figure 1.

**Figure 1:**
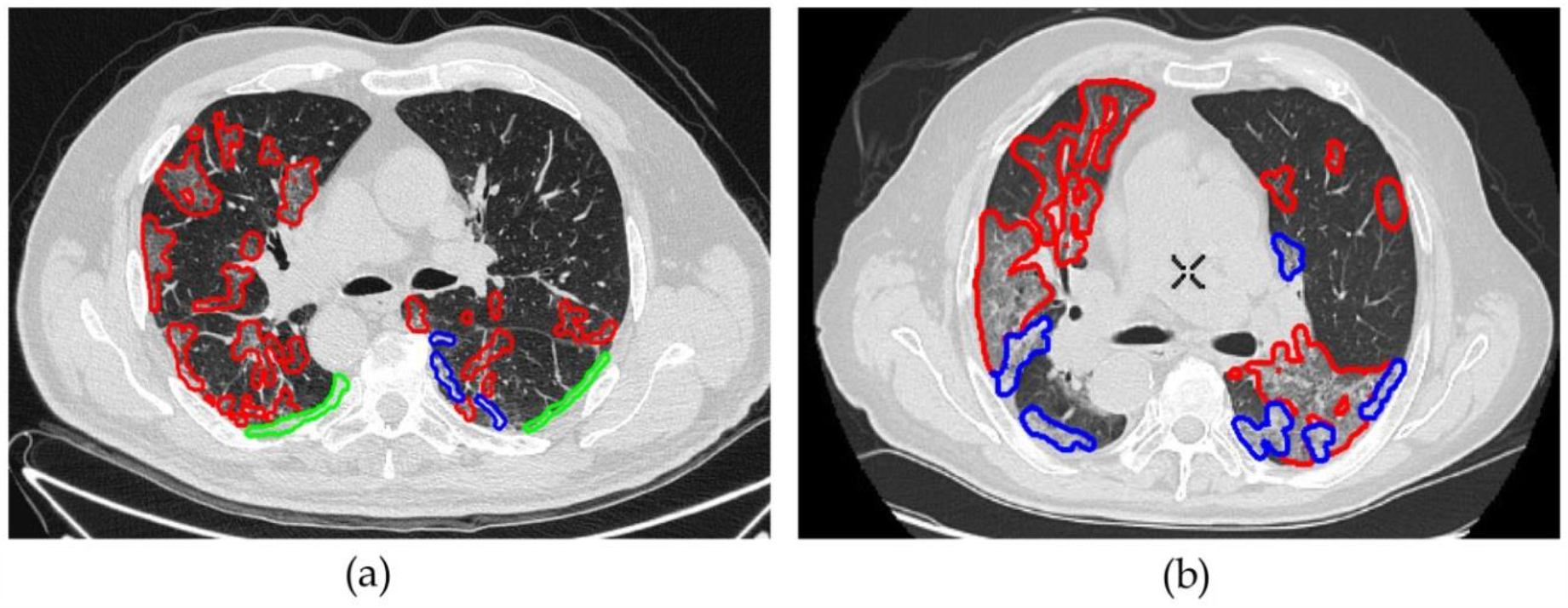
(a) One CT image slice with three distinctive regions – GCO (red), consolidation (blue) and pleural effusion (green). (b) two distinctive regions GCO (red) and consolidation (blue).

Radiomics features extracted from the whole lung provides the combinatorial outcome of the features of each of the three different regions. On the other hand, not all the features are able to distinguish between regions and are suitable to be used as image based biomarkers for accurate diagnosis, evaluation as well as monitoring disease progression and response to therapy. Since GCOs have been shown to precede consolidations, it is vital to determine the most suitable radiomics features that can distinguish and quantify different regions. This requires detail and careful investigation. According to the knowledge of the author, no such study has been conducted for COVID-19 lung CT regions. This study aims to address these by determining the radiomics features that can characterize each of the regions separately and can distinguish the regions from each other.

## MATERIALS AND METHODS

COVID-19 CT images were collected from available data derived from Italian Society of Medical and Interventional Radiology website [20, 21]. A total of 100 CT slices of 43 COVID-19 patients were available. GCO, consolidation and pleural effusion were segmented by a radiologists. Out of 100 slices, 96 slices have GCO, 78 slices have consolidation and 25 slices have pleural effusion. Only 23 of slices of 17 patients have all three regions. Rest of the slices had any one or two regions. A total of 44 radiomics features were extracted as shown in Table 1. Out of these 44 radiomics features, 43 features are textural features with the first three features are global features extracted from image histogram [22]. These 43 features can be broadly categorized into six different groups-1) global texture based on histogram, 2) Grey Level Co-Occurrence Matrix (GLCM), 3) Grey Level Run Length Matrix (GLRLM), 4) Grey Level Size Zone Matrix (GLSZE) and 5) NGTDM. To extract the rest 40 features, each region needs to be quantized to remove dependency on the image intensity.

**Table 1:**
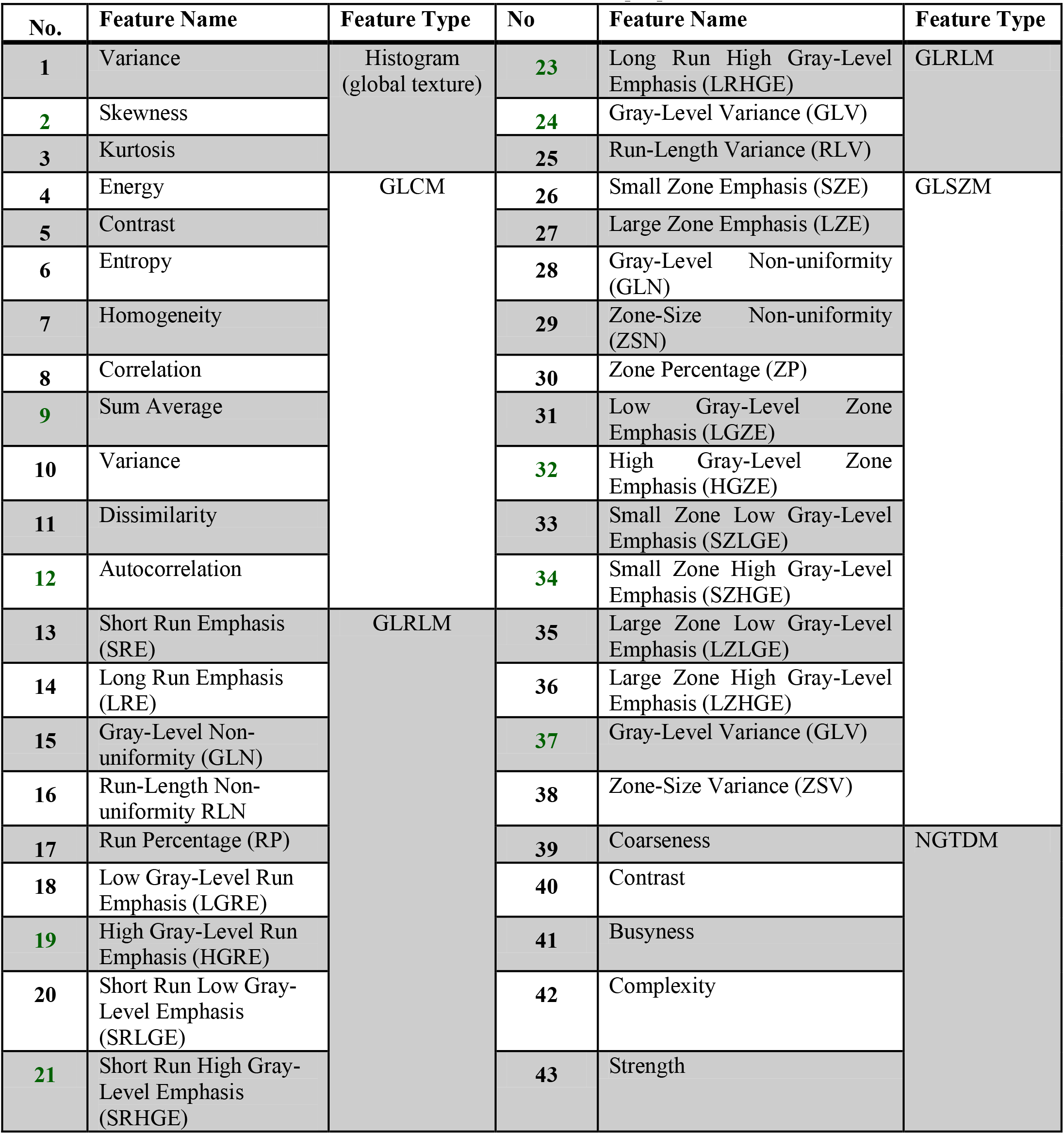
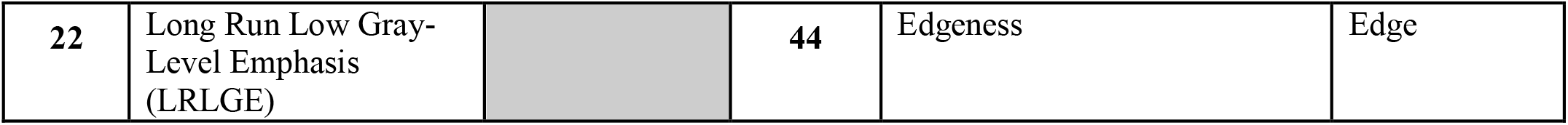
Radiomics Features [22]

To investigate if quantization has any impact on the features, four different levels of quantization were used (16, 32, 64 and 128). The final feature is an edge feature where numbers of pixels having edges more than a certain threshold within a region were counted.

Two approaches were implemented to determine the features that can differentiate between lung pathological regions. Firstly, Z-score and correlation heatmaps were generated for all regions for all 100 slices to identify the features that can classify different regions and to remove redundant features respectively. Secondly, one way ANOVA was then performed only on 23 slices that contain all the three regions to determine the features that are statistically significantly different () between the regions. Radiomics features that show agreement for all cases (Z-score, correlation and statistical significant test) were selected as suitable features and were applied on separate 52 slices containing only GGO and consolidation to test their accuracies by evaluating their *p-*values.

## RESULTS

Figure 2 shows the Z-score heatmap of all 44 features. The Z-score for each patient were calculated and arranged according to different regions. The correlation coefficients along with the *p*-values for testing the hypothesis of no correlation for all the 44 features are shown in Figure 3. Careful investigation of Figure 2 reveals that 10 radiomics features have better classification ability between GGO, consolidation and pleural effusion. These features are also have lower correlation (Figure 3).

**Figure 2:**
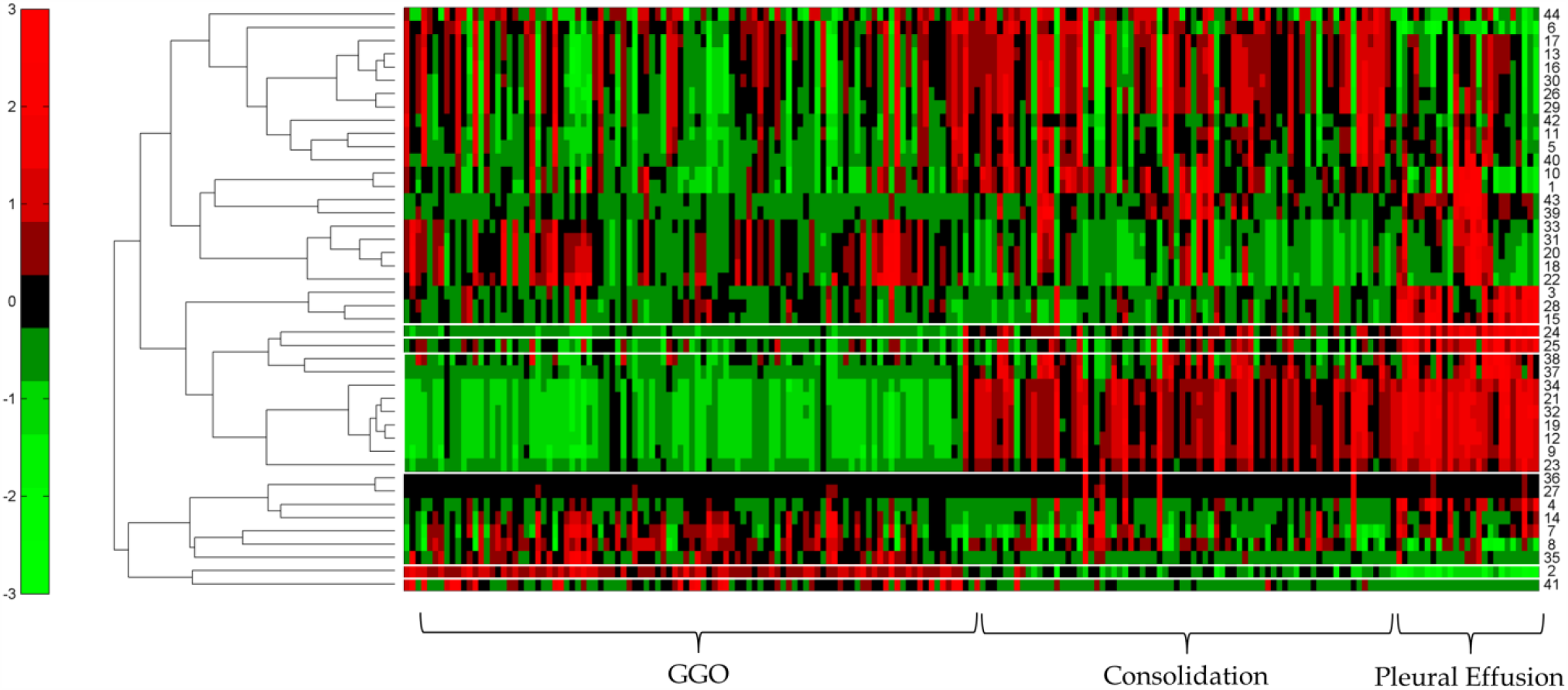
Z-score heatmap of all 44 features. Each row corresponds to one feature represented by one number (Table 1). Each column represent one regions. Left 96 column represent GGO, middle 78 represent consolidation and last 25 represent pleural effusion. The features that can distinguish between these three pathological regions are shown by the white bounding box.

**Figure 3:**
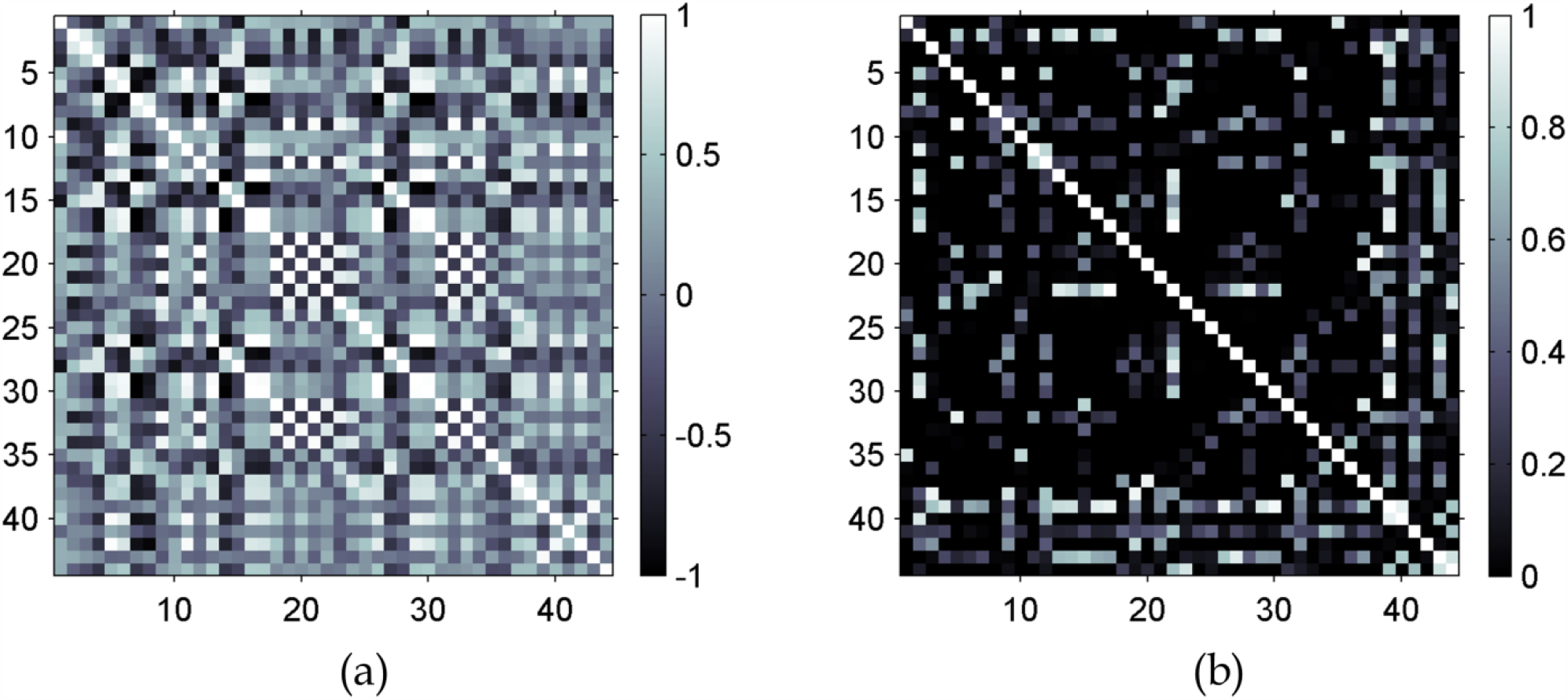
(a) correlation coefficient of 44 radiomics features. (b) *p* values describing the relationship between the features.

Out of 44 radiomics features, the same 10 features were found to be statistically significantly different between GCO, consolidation and plural effusion and do not depend on the quantization level. The features are shown in Table 2 along with their corresponding *p* values for three different regions.

**Table 2:**
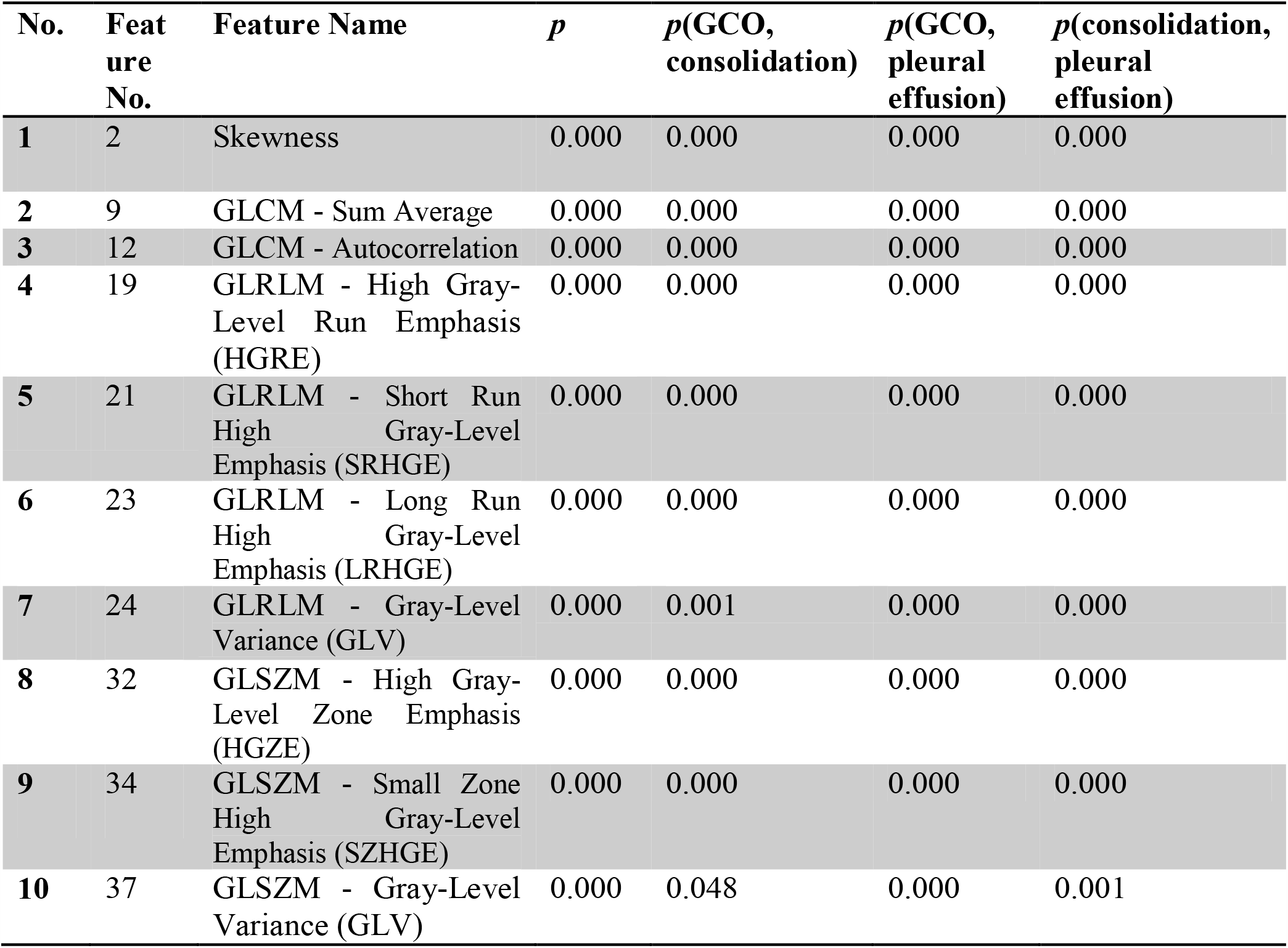
*p*-values extracted with one way ANOVA

The corresponding boxplots of all the three regions for 10 different radiomics features are shown in Figure 4.

**Figure 4:**
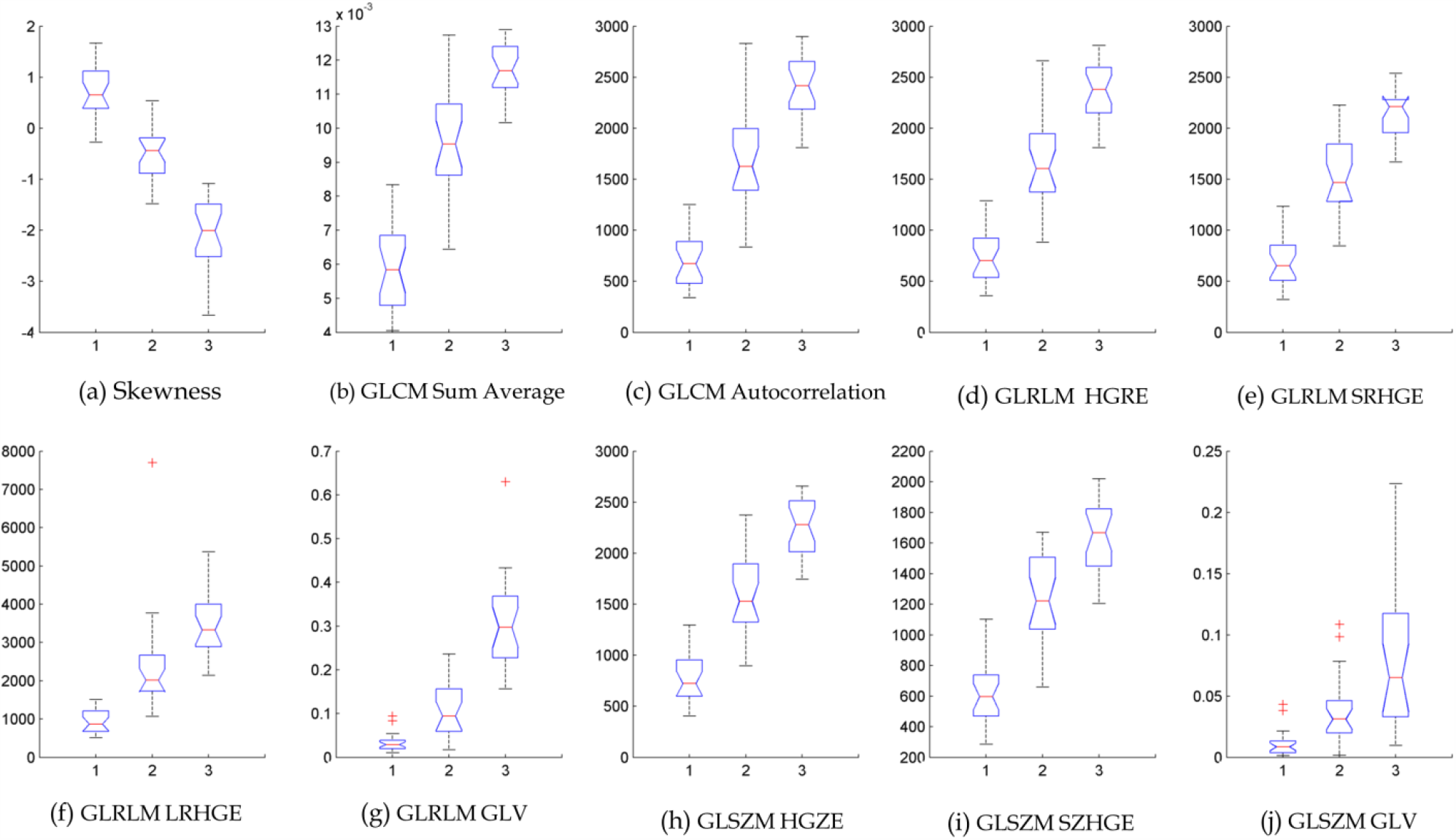
Box plot of 23 slices (notches in the boxplot representing group medians) of 10 radiomics features that are significantly different for three different regions (1- GGO, 2- Consolidation and 3- Pleural Effusion).

These 10 radiomics features (Table 2) for GGO and consolidation for the other 52 slices as shown in Figure 5. All these features were statistically significantly different between the two regions with the corresponding *p*-values were always less than 0.05.

**Figure 5:**
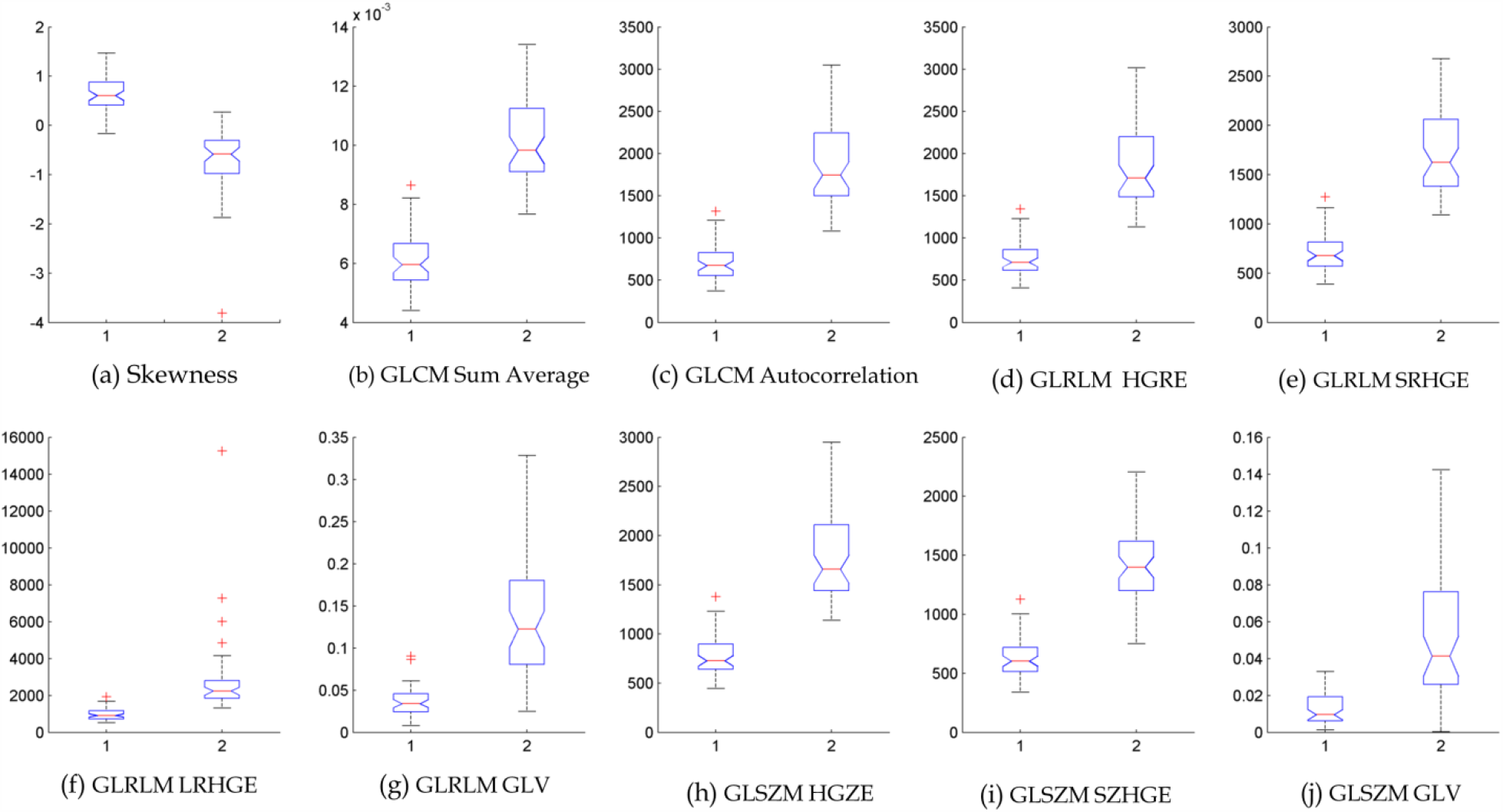
Box plot of 52 slices (notches in the boxplot representing group medians) of 10 radiomics features for GGO and consolidation (represented by 1 and 2 on the X-axis respectively).

## DISCUSSION

Because of its high sensitivity specially within the first two days of infection and capability to provide detail condition of the patient, computed tomography (CT) is routinely being used as a diagnostic and screening tool in several countries [23]. It also shows earlier positive sign of improvement for those patients who recover compared to RT-PCR results [23, 24].

The CT images of COVID-19 patients show distinctive patterns of infection with the hallmarks being bilateral and peripheral ground-glass and consolidation [25]. Sometimes CT examination showed a mixed and diverse pattern with the involvement of both lung parenchyma and interstitium [26] that includes bilateral multilobar ground-glass opacification (GGO) with a peripheral or posterior distribution [27]. It has been reported that temporal changes in CT manifestations can indicate progression and recovery of COVID-19 [28]. A manual chest CT severity score (CT-SS) has been proposed for COVID-19 to rapidly identify patients with severe forms of the disease [29]. 83.3% sensitivity and 94% specificity were achieved with this approach. Lung abnormalities on chest CT scans reported to show greatest severity approximately 10 days after initial onset of symptoms [30].

All these studies were performed retrospectively where radiologist had the time to examine each scan in detail. However, with the increasing number of CT scans performed on COVID-19 patients, it becomes difficult for the radiologists to examine the scan accurately in real time. On the other hand, intra- and inter-observer variability can introduce uncertainty in the staging the disease. Moreover, all the textural pattern that are considered to be the hallmark of COVID-19 may not be visible to the naked human eye especially it becomes more challenging in 3D.

Because of these reason, there is a dire need of a CT diagnostic method that can perform the task of rapid and early detection and monitoring quantitatively with high repeatability. Radiomics features can provide quantitative information regarding tissue heterogeneity and have been correlated with disease progression and prognosis in cancer.

In contrast to the previous radiomics studies where the focus was to extract the radiomics features for the whole lung of the COVID-19 patient [18, 19], this study focused on the regional extraction of the radiomics features from CT images. The argument for such approach is that when radiomics features are extracted for whole lung the regional information is diluted that can carry vital information regarding the disease stage, progression and response to therapy. So it was important to find the radiomics features that can distinguish different regions.

This investigation confirms that not all the proposed radiomics features can distinguish between GGO, consolidation and pleural effusion. Only a handful of features are able to differentiate between these regions and can be used for accurate quantitation of the disease.

## CONCLUSION

To the best of author’s knowledge, this paper presents the first study to determine radiomics features that can differentiate between ground glass opacity, consolidation and pleural effusion from CT data for COVID-19 patients. The study reveals that only ten radiomics features can differentiate between these regions. They are – 1) Skewness, GLCM – Sum Average, 3) GLCM –Autocorrelation, 4) GLRLM - High Gray-Level Run Emphasis (HGRE), 5) GLRLM - Short Run High Gray-Level Emphasis (SRHGE), 6) GLRLM - Long Run High Gray-Level Emphasis (LRHGE), 7) GLRLM - Gray-Level Variance (GLV), 8) GLSZM - High Gray-Level Zone Emphasis (HGZE), 9) GLSZM - Small Zone High Gray-Level Emphasis (SZHGE) and 10) GLSZM - Gray-Level Variance (GLV). These ten features can be useful in extracting quantitative data from CT lung images to characterize the disease in the patient, which in turn can help in more accurate diagnosis, staging the severity of the disease and allow the clinician to plan for more successful personalized treatment for COVID-19 patients. Future work is planned to link these quantitative radiomics information with the clinical outcomes so that they can be used for monitoring the progression of COVID-19 and response to therapy for clinical trials.

## Data Availability

The data used in this study are available.

https://medicalsegmentation.com/covid19/

